# Genome-wide association study of enlarged perivascular spaces: results from two large population-based cohorts

**DOI:** 10.1101/2025.09.09.25335402

**Authors:** Deborah Früh, Aslam Mohammed Imtiaz, Santiago Estrada, Mohammad Shahid, N. Ahmad Aziz, Monique M.B. Breteler

**Affiliations:** Population Health Sciences, German Center for Neurodegenerative Diseases (DZNE), Bonn, Germany; Artificial Intelligence in Medical Imaging, German Center for Neurodegenerative diseases (DZNE), Bonn, Germany; Department of Neurology, Faculty of Medicine, University of Bonn, Germany; Institute for Medical Biometry, Informatics and Epidemiology (IMBIE), Faculty of Medicine, University of Bonn, Germany

**Author notes:** Corresponding author: Prof. Dr. Dr. Monique M. B. Breteler Population Health Sciences, German Center for Neurodegenerative Diseases (DZNE) Venusberg-Campus 1/99, 53127 Bonn Germany.

## Abstract

Enlarged perivascular spaces (PVS) are markers of cerebral small vessel disease (cSVD), implicated in neurovascular and neurodegenerative processes. We conducted a genome-wide association study (GWAS) of quantitative, global PVS volume using brain MRI data from UK Biobank and the Rhineland Study. The meta-analysis (n = 43,050) identified 54 genome-wide significant loci, including 39 novel loci, implicating vascular, inflammatory, cytoskeletal, and metabolic pathways. A parallel GWAS of white matter hyperintensity volume in the same sample showed no significant genetic correlation with PVS, suggesting distinct genetic architectures. Mendelian randomization supported a robust causal effect of systolic and diastolic blood pressure on PVS volume, independent of hypertension diagnosis. We also observed suggestive causal evidence for plasminogen activator inhibitor-1, indicating a potential role for prothrombotic and inflammatory processes. These findings reveal a multifactorial genetic architecture for PVS and underscore vascular dysfunction as a central contributor to cSVD, with implications for prevention and risk stratification.

## Introduction

Cerebral small vessel disease (cSVD) is an umbrella term for a group of prevalent and progressive neuropathologic processes that affect the cerebral microstructure of the white matter (WM) and deep grey matter structures.^1^ As one of the leading causes of stroke, cognitive decline and dementia, cSVD represents a critical area of research and a key target for preventing these conditions.^1–3^ Beyond these clinical outcomes, cSVD is also linked to gait disturbances, mood disorders, and sleep disruption, though it may remain asymptomatic for years.^2,4^ The underlying pathology of cSVD is closely tied to vascular risk factors such as hypertension, diabetes mellitus, and smoking, among others.^1,2,5^

Several magnetic resonance imaging (MRI)-based biomarkers of cSVD have been identified, including white matter hyperintensities (WMH), enlarged perivascular spaces (PVS), recent small subcortical infarcts, lacunes, cerebral microbleeds, cortical superficial siderosis and brain atrophy.^4,6^ These neuroradiological markers of cSVD are common and become nearly ubiquitous with increasing age. However, early signs of cSVD, particularly WMH and PVS, are also identifiable in younger individuals, suggesting that vascular changes begin early in life and progress over decades.^6,7^

Among the cSVD imaging biomarkers, WMH are studied the most. In contrast, PVS, also known as Virchow-Robin spaces, have only recently gained more attention. PVS are cerebrospinal fluid (CSF)-filled cavities, surrounding small penetrating vessels, like capillaries, arterioles, and venules and are integral to the brain’s waste clearance system.^8,9^ Only when enlarged, they become visible on MRI scans and are considered to reflect perivascular dysfunction and possible impairment of the brain’s waste management.^6,10^ PVS are proposed to be among the earliest detectable signs of cSVD.^11^ Mechanisms underlying PVS dilation are thought to be heterogeneous, including PVS obstructions due to neuroinflammation, oxidative stress and vascular stiffening, impairment of interstitial fluid circulation, spiral elongation of arteries and brain atrophy or myelin loss.^12,13^ Despite growing interest, however, these mechanisms remain incompletely understood.

Given the clinical and biological importance of cSVD, elucidating its genetic underpinnings has emerged as a powerful approach for uncovering disease mechanisms. Genome-wide association studies (GWAS) have identified multiple loci associated with WMH volume with reported heritability estimates ranging from 49% to 80%, highlighting a substantial genetic component.^5,14–16^ On the contrary, genetic studies on PVS remain comparatively limited, largely due to challenges in obtaining accurate measures of PVS volumes. Given the high prevalence and widespread distribution of PVS throughout the whole brain, manual rating or tracing is labor-intensive and thus unfeasible for large cohorts and automated segmentation approaches require precise manual annotations for effective training.^17,18^ Consequently, the only GWAS to date, which revealed 24 genome-wide significant PVS burden risk loci, was based on semiquantitative visual ratings in selected brain regions, i.e., white matter, basal ganglia (BG) and hippocampus.^11^ While providing valuable insights into the biology of PVS and pointing to pathways involved in PVS etiology and progression, this approach for quantifying PVS may have limited the detection of genetic signals related to more nuanced or widespread PVS involvement. Recent advances in artificial intelligence-based computational methods for PVS quantification allow to capture PVS in a more fine-grained manner and offer the opportunity to assess them on a continuous spectrum as well as within the whole brain.

Combining data from two large population-based cohorts, i.e., the Rhineland Study and UK Biobank, we performed a GWAS using continuous, quantitative measures of PVS volume, rather than categorical or visual burden scores, capturing subtle variation across individuals. Furthermore, we assessed whole-brain PVS volume, rather than restricting analyses to a few specific regions. Finally, we directly compared the genetic architecture of PVS and WMH to assess the extent of genetic overlap between two distinct markers of cSVD. By leveraging recent advances in automated segmentation pipelines of PVS, we aim to provide a more comprehensive understanding of the genetic basis of cSVD. Our results uncover novel loci associated with PVS volume, not identified in the prior region-specific and semiquantitative study.

## Materials and Methods

### Study population

The Rhineland Study (RS) is an ongoing, prospective, population-based cohort study in Bonn, Germany, enrolling participants aged 30 years and older from two districts. The only exclusion criterion is insufficient German language proficiency to provide informed consent. Participants undergo comprehensive deep phenotyping, including questionnaires, bio-sample collection, neurological exams, and a one-hour brain MRI scan session. The study was approved by the Ethics Committee of the University of Bonn Medical Faculty (Ref: 338/15) and follows ICH-GCP standards, with written informed consent in accordance with the Declaration of Helsinki.

The UK Biobank (UKB) is a large-scale, population-based cohort study of ∼500,000 individuals aged 40-69 years at baseline from across the UK. It offers extensive health data, including bio-samples, genetic information, and questionnaires. A subset of participants underwent additional imaging, including brain MRI. The study was approved by the NHS North West Research Ethics Committee (Ref: 11/NW/0382), with written informed consent obtained from all participants.

Our GWAS was based on 5838 participants from the Rhineland Study with imaging data and 46569 participants from the UKB imaging sub-study. After the exclusion of participants due to insufficient image quality or failed MRI post-processing (Rhineland Study: n = 107; UKB: n = 1579) and missing genetic data, ethnic outliers and sex aneuploidy outliers (Rhineland Study: n = 686; UKB: n = 6985), our analytic sample consisted of 5045 participants from the Rhineland Study and 38005 participants from the UKB.

### Brain MRI

MRI in the Rhineland Study was performed on two equal 3T MRI scanners (MAGNETOM Prisma, Siemens Healthineers) a 64-channel head-neck receive coil. For segmentations of cSVD biomarkers T1- weighted (0.8 mm isotropic image resolution), T2-weighted (0.8 mm isotropic) and T2 fluid-attenuated inversion recovery (FLAIR, 1.0 mm isotropic) structural MR images were used.^19^ UKB imaging data was acquired on Siemens Skyra 3T scanners equipped with 32-channel head coils at three sites.^20^ Detailed information on imaging parameters in UKB are available in the online documentation (biobank.ctsu.ox.ac.uk/crystal/crystal/docs/brain_mri.pdf).

Segmentation maps of WMH in the Rhineland Study were automatically generated with an in-house developed pipeline^21^, based on DeepMedic^22^, using multi-modal MRI as input. In UKB total volume of WMH were calculated using BIANCA^23^ utilizing both T1-weighted and T2-FLAIR weighted images.^24^ We quantified volumes of enlarged PVS using an automated segmentation pipeline based on deep learning, adapted from a previous study^25^ and retrained with additional manual labels primarily from the Rhineland Study cohort to improve the tools generalizability. Based on the requirements of the PVS pipeline, Rhineland Study images were down-sampled to 1mm isotropic resolution. Volumes of PVS and WMH were logarithmically transformed to correct for skewness. WM volumes were extracted using FreeSurfer’s automated segmentations in both cohorts (version 6.0).^26^ WMH load and PVS load were defined as the global WMH/PVS volume normalized to the global WM volume to account for varying WM volumes due to head size, atrophy or lesions.

### Additional Phenotypes

In addition to brain MRI endophenotypes, we considered several additional traits for descriptive statistics or secondary analysis. These included measures of blood pressure, hypertension status, smoking behavior, body mass index (BMI) and sleep duration.

Systolic and diastolic blood pressure (SBP and DBP) were measured with an automated oscillometric upper arm blood pressure monitor in both studies. If automatic measurements failed in UKB, manual measurements were performed. Measurements were averaged across two or more readings where available. Hypertension was defined as SBP ≥140 mmHg, DBP ≥90 mmHg, or current use of antihypertensive medication. Smoking behavior was self-reported and categorized as current smoker, former smoker, or never smoker. For certain analyses, smoking status was dichotomized as current versus non-current smoker. BMI was calculated as weight in kilograms divided by height in meters squared (kg/m²). Average self-reported sleep duration was obtained from questionnaire data and divided into a four-level score based on the Pittsburgh Sleep Quality Index component 3.

### Genetic Data

Genotyping in the Rhineland Study was performed with the Omni 2.5 Exome array, and genotypes were called from raw signal intensity files using GenomeStudio. In UKB, UK BiLEVE Axiom array and UK Biobank Axiom array were used for genotyping and genotype calling was performed by Affymetrix.^27^ Quality control of SNPs in both datasets was performed by excluding SNPs with missingness rates > 98%, minor allele frequencies < 1% (if palindromic >0.4%) and deviation from the Hardy-Weinberg Equilibrium (p-value < 10^-6^). Quality control of samples was performed separately by excluding all samples with missingness rates >95%, abnormal heterozygosity and ethnic outliers as described previously.^28^ After quality control, missing genetic data was imputed using Impute2^29^ with 1000 Genomes (version 3, phase 5) and HRC as the reference panel in Rhineland Study ^30^ and UKB^31^, respectively. SNPs with an imputation score <0.3 were excluded.

### Genome-wide association analysis

To conduct the GWAS analyses, we used REGENIE (v2.2).^32^ REGENIE applies a two-step approach for genome-wide association analysis. In Step 1, a whole-genome regression model is fitted using a subset of genotyped variants to capture the polygenic contribution to phenotypic variance. Step 2 then tests the association of imputed SNPs with the phenotype, conditioning on predictions from Step 1 and employing a leave-one-chromosome-out scheme to avoid proximal contamination. In Step 1, genotyped SNPs were pruned based on linkage disequilibrium (LD). For the UKB, in Step 1 of REGENIE, pruning was performed with a threshold of r² < 0.25, a window size of 200 variants, and a step size of 50 to reduce redundancy among highly correlated SNPs, minimize the risk of overfitting the whole-genome regression model, and improve computational efficiency. For the Rhineland Study, given its smaller sample size, we used an LD threshold of r² < 0.9 with a window size of 1,000 variants and a step size of 100 to retain more variants to enable better capturing of the polygenic signal. The GWAS was performed on log-transformed PVS/WMH volumes assuming additive genetic effects in two models. Our base model (termed model 1) adjusted for age, sex, ETIV, and the first 10 genetic principal components. The second model (termed model 2) additionally considered the hypertension status of participants. A fixed-effects inverse variance-weighted meta-analysis was conducted using METAL^33^, based on the GWAS summary statistics from both models in Rhineland Study and UKB.

As a sensitivity analysis, we conducted binary case-control GWAS using categorized participant groups based on PVS burden. Individuals in the lower 10th percentile of the distribution for PVS volume (n = 505) were classified as the PVS-absence group, while control groups consisted of individuals in either the upper half or the interquartile range of the distribution (n = 2522). Given the strong age dependence of PVS volumes, we also applied an alternative group definition, selecting the lower 10th percentile within each 10-year age stratum (n = 509).

To assess the potential influence of different inheritance patterns, we conducted additional sensitivity analyses testing recessive effects by assuming effects only in individuals homozygous for the minor allele using REGENIE.

Additionally, we tested for SNP-by-sex interaction effects on the phenotype by including a SNP × sex interaction term using the interaction option in REGENIE.

### Risk loci definition

To identify genomic risk loci, we used the Functional Mapping and Analysis of GWAS (FUMA) platform.^34^ They were defined as significant SNPs with LD scores r^2^ ≥ 0.6. LD blocks of independently significant SNPs within 250 kb of each other were joined into a single genomic locus. The lead SNP of a locus was defined as SNPs that were independent of one another at r^2^ < 0.1.

### Gene mapping

SNPs were positionally mapped to genes based on physical proximity (within 10 kb) using ANNOVAR.^35^ To further explore the biological context, we used the GENE2FUNC module in FUMA to assess the enrichment of the mapped genes in the GWAS catalog database of reported genetic variant-trait associations. Statistical significance was defined using the default Bonferroni-adjusted p-value threshold of < 0.05, with the additional requirement that each enriched gene set has to include at least two overlapping input genes.

To investigate functional consequences of associated variants, we performed expression quantitative trait loci (eQTL) mapping using expression data from brain and vascular tissues, drawing specifically on GTEx v8^36^, BRAINEAC^37^, PsychENCODE^38^, and the Common Mind Consortium^39^. Chromatin interaction mapping was conducted using FUMA, which integrates three-dimensional DNA-DNA interaction data to link PVS-associated loci with putative target genes. This included significant chromatin interactions from PsychENCODE enhancer-promoter links^38^ and promoter-anchored loops^39^, as well as HiC datasets from relevant brain regions such as adult and fetal cortex^40^, dorsolateral prefrontal cortex, and hippocampus, alongside data from neural progenitor cell lines.^41^ In addition, we mapped genes that overlapped with predicted enhancer regions (±250 bp upstream and 500 bp downstream of transcription start sites) using epigenomic annotations from 17 brain-related datasets provided by the Roadmap Epigenomics Project (i.e., E053, E054, E067, E068, E069, E070, E071, E072, E073, E074, E081, E082, E003, E008, E007, E009, E010, E057).^42^

### Genetic correlation analysis

We used LD score regression (LDSC)^43^ using GWAS summary statistics to estimate SNP heritability and the genetic correlation of PVS and WMH volume with regional PVS burden and Diffusion Tensor Imaging - Along the Perivascular Spaces (DTI-ALPS)^44^ as an alternative marker of glymphatic function. We additionally assessed genetic correlations with vascular risk factors (i.e., waist-to-hip ratio^45^, BMI^45^, alcohol consumption, SBP^46^, DBP^46^, high-density lipoprotein (HDL) cholesterol^47^, low-density lipoprotein (LDL) cholesterol^48^, type 2 diabetes^49^ and plasminogen activator inhibitor-1 (PAI-1)^50^), as well as neurodegenerative diseases and neurological traits (i.e., stroke, small vessel stroke^51^, Parkinson’s disease^52^, Alzheimer’s disease^53^ and educational attainment^54^), sleep related traits (i.e., chronotype^55^, daytime napping^56^ and sleep duration^56^) and blood based biomarkers of neurodegenerative diseases (amyloid-beta 40 (Aβ-40), amyloid-beta 42 (Aβ-42), their ratio (Aβ 40/42)^57^ and neurofilament light chain levels (NfL)^58^) (Supplementary Table 1). We used HapMap3 SNPs^59^ to calculate the genetic correlations and corrections for multiple testing were performed using the false discovery rate (FDR) method.

### Two-sample Mendelian randomization

To test if PVS volume was causally related to vascular risk factors or neurological outcomes we performed two-sample Mendelian randomization (MR). To minimize bias due to sample overlap, we selected independent GWAS summary statistics for Alzheimer’s disease, Parkinson’s disease, stroke, and small vessel stroke that did not include UKB participants. Because publicly available GWAS data for SBP and DBP include UKB data, we conducted a separate GWAS for SBP and DBP within the UKB, explicitly excluding individuals who were part of our primary PVS GWAS. This approach was implemented to mitigate potential overfitting and bias in the MR analyses. MR analyses were subsequently performed using these subset-derived instruments to check whether observed associations with PVS volume remained consistent. We used the inverse variance weighted (IVW)^60^ regression method as the primary test method, complemented by other methods (including MR Egger regression^61^, and weighted median, simple mode and weighted mode estimators^62^) for sensitivity analysis. All p-values were corrected for multiple testing using FDR. We evaluated heterogeneity in the IVW and MR-Egger estimates with Cochran’s Q-test.^63^ When heterogeneity was detected, radial MR^64^ was applied to identify outlier instrumental variables. Using the same framework, we also conducted reverse causation analyses. To assess potential weak instrument bias, we calculated the F-statistic, with values below 10 indicating weak genetic instruments.^65,66^

## Results

### Sample characteristics

Our GWAS meta-analysis was based on 43050 participants from two population-based cohorts. Rhineland Study participants had a mean age of 56.0 ± 13.6 years [range: 30.2 - 95.4 years] with 57.7% of the population being women. UKB participants were 64.5 ± 7.7 years [range: 45 - 83] on average and 52.4% were women. The mean PVS volume was 3709.9 ± 3145.2 mm^3^ in Rhineland Study and 1866.0 ± 1724.8 mm^3^ in UKB, while the mean count of PVS was 195.3 ± 105.2 in Rhineland Study and 105.8 ± 66.9 in UKB. More detailed sample characteristics can be found in Table 1.

**Table 1:** Sample characteristics in UK Biobank (UKB) and Rhineland Study (RS). WMH white matter hyperintensities, PVS enlarged perivascular spaces, eTIV estimated intracranial volume, WM white matter, BP blood pressure, BMI body mass index.

|  | UKB (n = 38005) | RS (n = 5045) |
| --- | --- | --- |
| Age [years] | 64.5 ± 7.7 | 56.0 ± 13.6 |
| Woman N (%) | 19920 (52.4) | 2909 (57.7) |
| Current smoking N (%) | 1170 (3.1) | 608 (12.1) |
| Alcohol consumption [g/kcal] | 1.95e-3 ± 2.50e-3 | 7.57e-3 ± 8.81e-3 |
| <u>MRI markers:</u> |  |  |
| WMH load [%] | 0.975 ± 1.27 | 0.422 ± 1.04 |
| PVS load [%] | 0.337 ± 0.298 | 0.817 ± 0.690 |
| PVS count | 105.8 ± 66.9 | 195.3 ± 105.2 |
| eTIV [cm3] | 1471.4 ± 158.9 | 1552.2 ± 151.9 |
| WM volume [cm3] | 545.4 ± 61.8 | 455.9 ± 58.7 |
| <u>Vascular risk factors:</u> |  |  |
| SBP [mmHg] | 139.7 ± 18.8 | 126.1 ± 15.6 |
| DBP [mmHg] | 78.9 ± 10.1 | 74.8 ± 9.11 |
| Hypertension N (%) | 20000 (52.6) | 1764 (35.0) |
| BMI [kg/m2] | 26.51 ± 4.38 | 25.6 ± 4.14 |
| <u>Sleep duration:</u> |  |  |
| > 7 hours N (%) | 13780 (36.3) | 1578 (31.1) |
| 6-7 hours N (%) | 14975 (39.4) | 2017 (40.0) |
| 5-6 hours N (%) | 8617 (22.7) | 1059 (21.0) |
| < 5 hours N (%) | 280 (0.7) | 341 (6.8) |

Phenotypic correlations with risk factors and clinical correlates are available in Supplementary Table 2. PVS load correlated most strongly with age (r = 0.41, P_FDR_ = < 2.2×10^-16^) and total arterial compliance index (r = -0.23, P_FDR_ = < 2.2×10^-16^) in Rhineland Study and age (r = 0.12, P_FDR_ = < 2.2×10^-16^) and WM volume (r = 0.16, P_FDR_ = < 2.2×10^-16^) in UKB. Correlation of PVS and WMH load was moderate in Rhineland Study (r = 0.19, P_FDR_ = < 2.2×10^-16^) and small in UKB (r = 0.05, P_FDR_ = < 2.2×10^-16^).

### Genome-wide association analysis

In the UKB discovery GWAS of global PVS volume, we identified 344 independent genome-wide significant SNPs in the base model (Supplementary Table 3, Supplementary Figure 1) and 295 in the hypertension-adjusted model (Supplementary Table 4, Supplementary Figure 1). Of these, 20 SNPs replicated in the Rhineland Study at a Bonferroni-corrected significance threshold of P < 0.05/n (Supplementary Figure 2).

The subsequent meta-analysis combining data from both UKB and Rhineland Study revealed 289 independently significant SNPs (Supplementary Table 5) across 50 genome-wide significant loci (P < 5×10^-8^) in the base model, and 254 SNPs (Supplementary Table 6) across 48 loci in the hypertension-adjusted model. Hypertension adjustment revealed two additional loci (11p11.12 near *RP11-574M7.2* and 19q13.42 near *TMEM150B*), while four base model loci no longer met genome-wide significance (Table 2, Figure 1). Thus, in total 54 loci reached genome-wide significance for PVS volume, of which 39 were not reported in previous GWAS. While our GWAS on continuous PVS measures replicated most of the findings from the earlier analysis of regional PVS burden, we did not replicate two previously reported risk loci for WM PVS burden (8p11.21 near *SMIM19*, *CHRNB3*; 11q13.3 near *FADD*, *PPFIA1*) and three loci previously reported for BG and hippocampal PVS (1q25.3 near *SHCBP1L*, *LAMC1*; 2q33.2 near *NBEAL1*, *ICA1L*; 3q26.31 near *TMEM212*). LD score regression-based heritability of global PVS volume was estimated at 29.8 ± 2.6% for the base model and 29.9 ± 2.7% for the hypertension-adjusted model.

The discovery GWAS of WMH volume in UKB identified 103 independent genome-wide significant SNPs, and 86 SNPs after adjusting for hypertension. Eight base-model SNPs and nine hypertension-adjusted SNPs replicated in the Rhineland Study (Supplementary Table 7-8). The following meta-analysis yielded 83 independent SNPs (Supplementary Table 9) corresponding to 20 genomic risk loci in the base model (Supplementary Table 10, Figure 1). The hypertension-adjusted model identified one additional locus (10q26.2 near *FANK1*) and one locus no longer met genome-wide significance (Supplementary Table 11). Overall, 68 SNPs were significantly associated with WMH volume in the adjusted model, corresponding to 20 loci.

**Figure 1:**
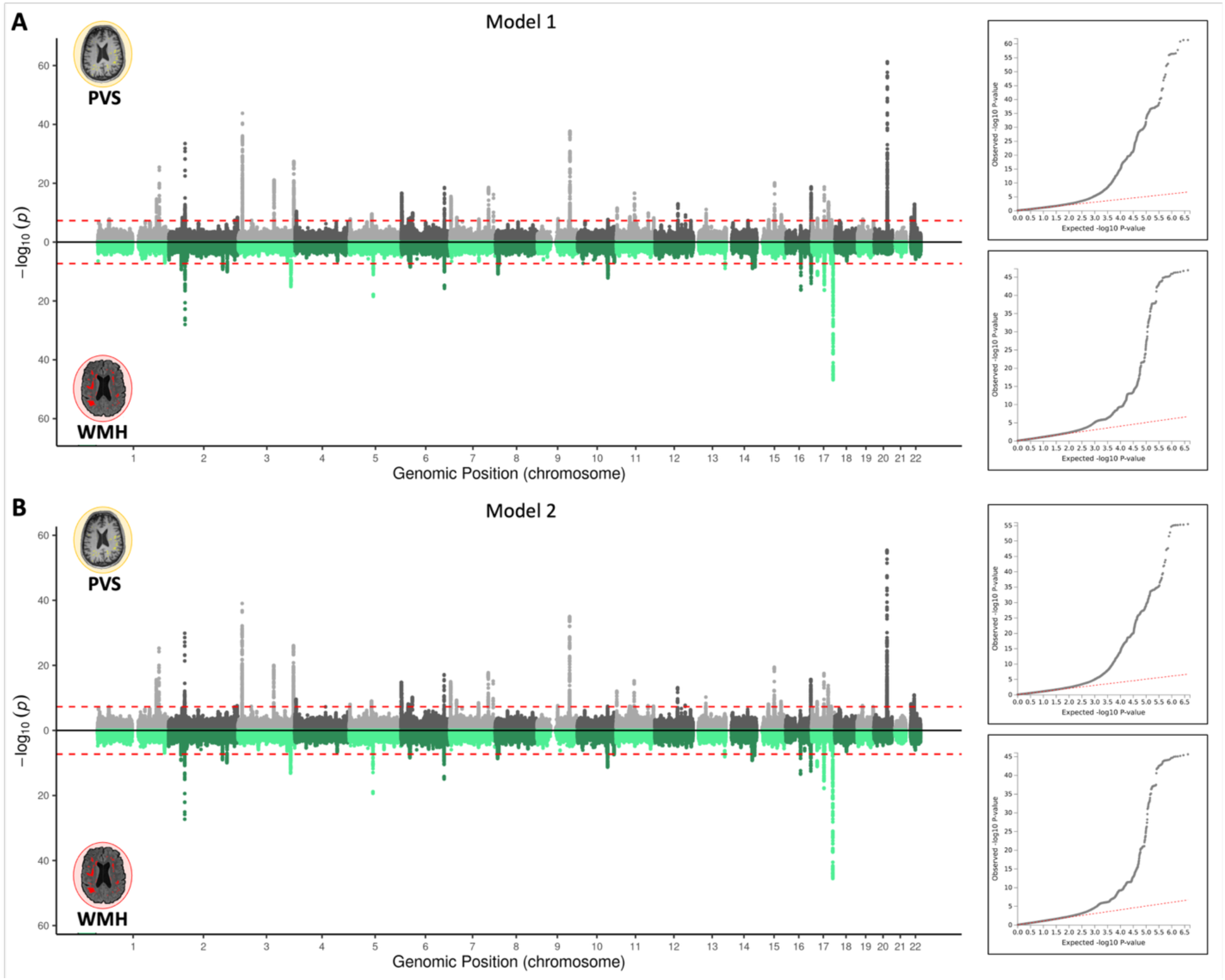
Mirror Manhattan plot of genome-wide association results for perivascular spaces (PVS) (top) and white matter hyperintensities (WMH) (bottom) in the meta-analysis of UK Biobank and Rhineland Study cohorts, with corresponding quantile-quantile (QQ) plots shown on the right. The top panel (A) shows results from the base model adjusted for age, sex, estimated total intracranial volume (eTIV), and genetic principal components; the bottom panel (B) shows results from model 2, which includes additional adjustment for hypertension.

**Table 2:**
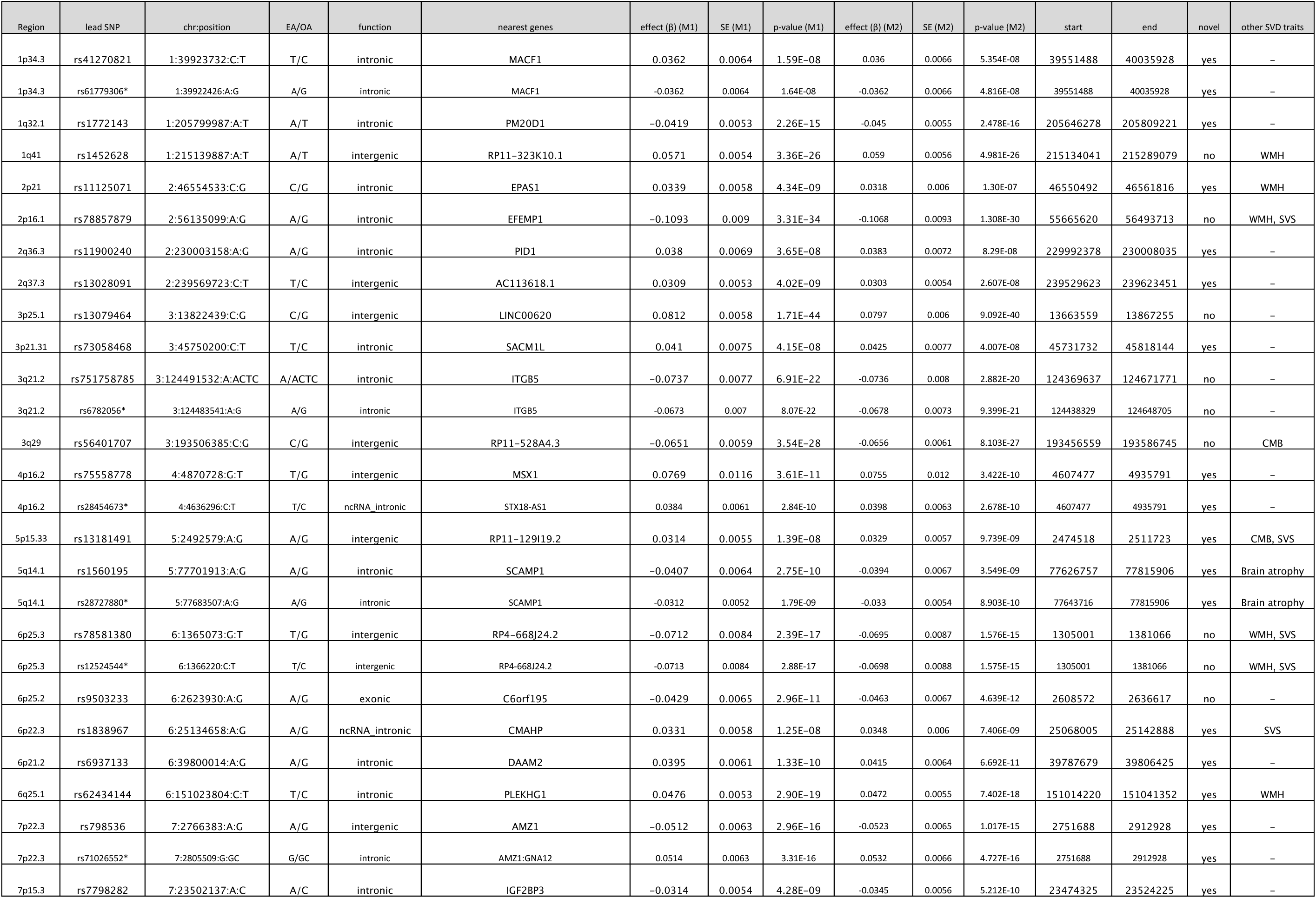
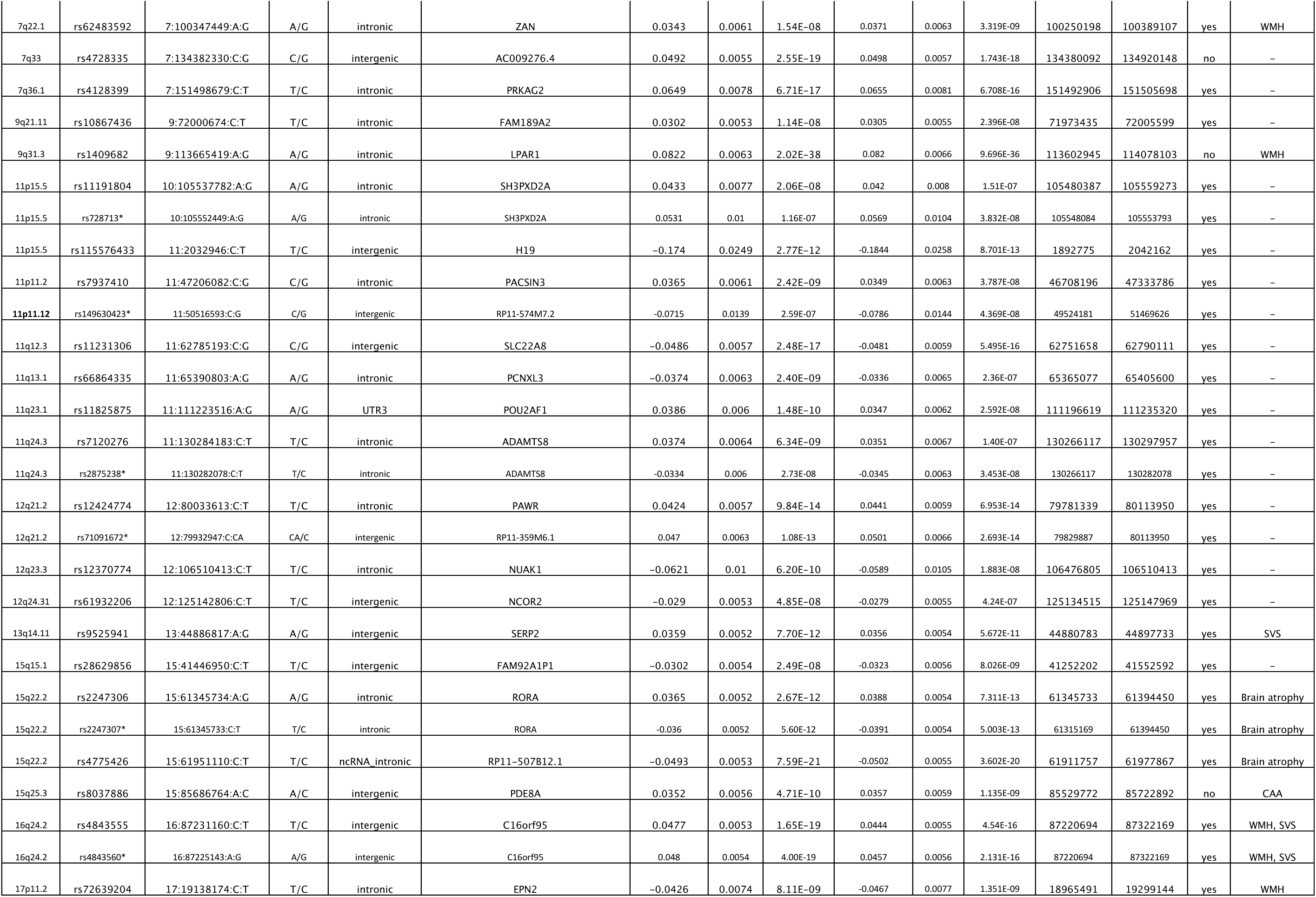
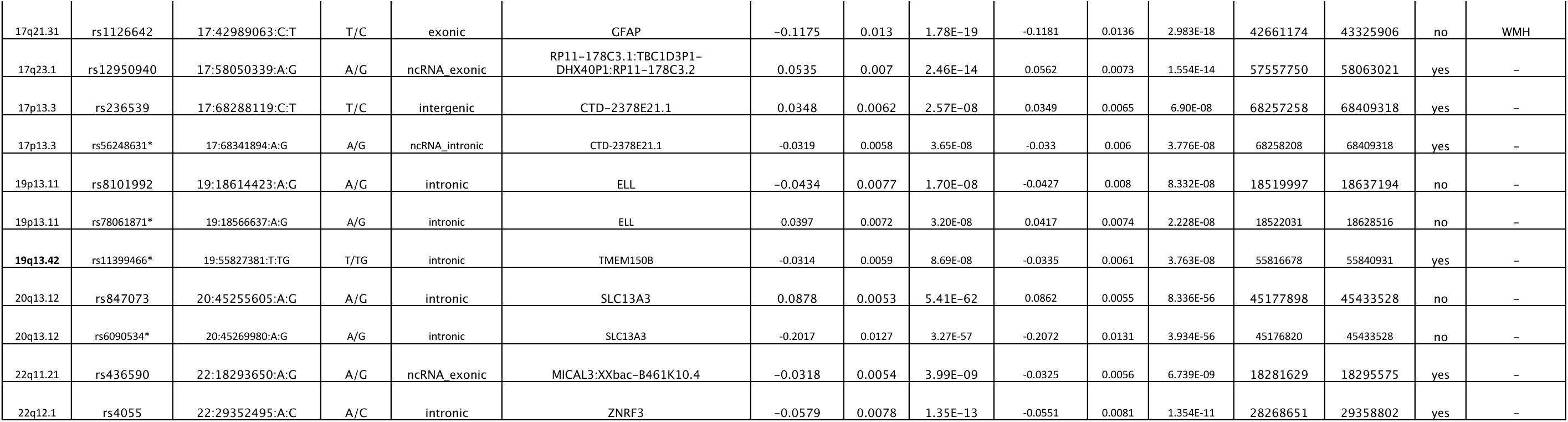
Genome wide significant loci for PVS trait from model 1 and model 2. Lead SNPs marked with * were different in model 2.; Novel column indicates if the genomic locus was identified previously for PVS trait. SNP single nucleotide polymorphism, chr chromosome, EA effect allele, OA other allele, β effect estimate, SE standard error, M1 model 1, M2 model 2.

To explore genetic factors contributing to disease resilience, we performed a supplementary GWAS comparing participants with PVS absence (lowest 10^th^ percentile) to controls (upper half or interquartile range of the distribution; Supplementary Table 12). This case-control approach identified several genome-wide significant loci, including some not reported in previous PVS burden GWAS (Supplementary Tables 13-14). However, most loci overlapped with those identified in the main quantitative PVS GWAS. Notably, one distinct signal (1q25.3 near *LAMC1*) emerged in the hypertension-adjusted, age-stratified model using the upper half of the distribution as controls. This locus had previously been linked only to hippocampal PVS burden (Supplementary Table 14).

In an additional analysis applying a recessive genetic model, we found that the recessive GWAS largely recapitulated the signals observed in the additive model for both PVS (Supplementary Table 15-16, Supplementary Figure 4) and WMH (Supplementary Table 17-18, Supplementary Figure 4), while no genome-wide significant loci were identified using a dominant model. However, two loci emerged in each of the base and hypertension-adjusted recessive GWAS models on PVS that were not detected under the additive model. Notably, one of these loci was 11q13.3 near *PPFIA1*, which was identified in both recessive models and has previously been associated with WM PVS burden.

To evaluate sex-specific genetic effects, we conducted sex-interaction analyses and stratified GWAS analyses in men and women separately (Supplementary Table 19). We identified one genomic risk locus represented by the lead SNP rs9877716 near *CHL1*, where the interaction term was statistically significant (β = -0.0597, P = 3.56 × 10⁻⁸), indicating differential SNP effects by sex. Stratified analyses showed nominal association, with stronger effect in women (β = -0.0350, P = 1.25 × 10⁻⁵) compared to men (β = 0.0236, P = 0.0036), consistent with the negative interaction coefficient (Supplementary Figure 5).

### Gene mapping and genetic correlation of biomarkers of cSVD

For each of the independently significant SNPs in both models and traits, we identified the nearest genes based on their physical proximity to the SNPs. We identified 117 different genes associated with PVS volume and 52 genes associated with WMH volume. Six of these were identified in both our PVS and WMH GWAS meta-analyses, indicating overlap between the two traits. Additionally, 12 genes uniquely associated with one trait in our analyses, had previously been reported in GWAS of the respective other trait (Figure 2). Furthermore, we looked up associations of the 54 PVS risk loci defined by the genomic position with other markers of cSVD i.e. WMH, cerebral microbleeds, small vessel stroke, brain atrophy and ventricular enlargement. We found that 22 out of 54 genomic risk loci (41%) showed significant association with at least one other cSVD trait. WMH volume showed the largest number of significant associations (12 loci) with PVS risk loci (Table 2).

**Figure 2:**
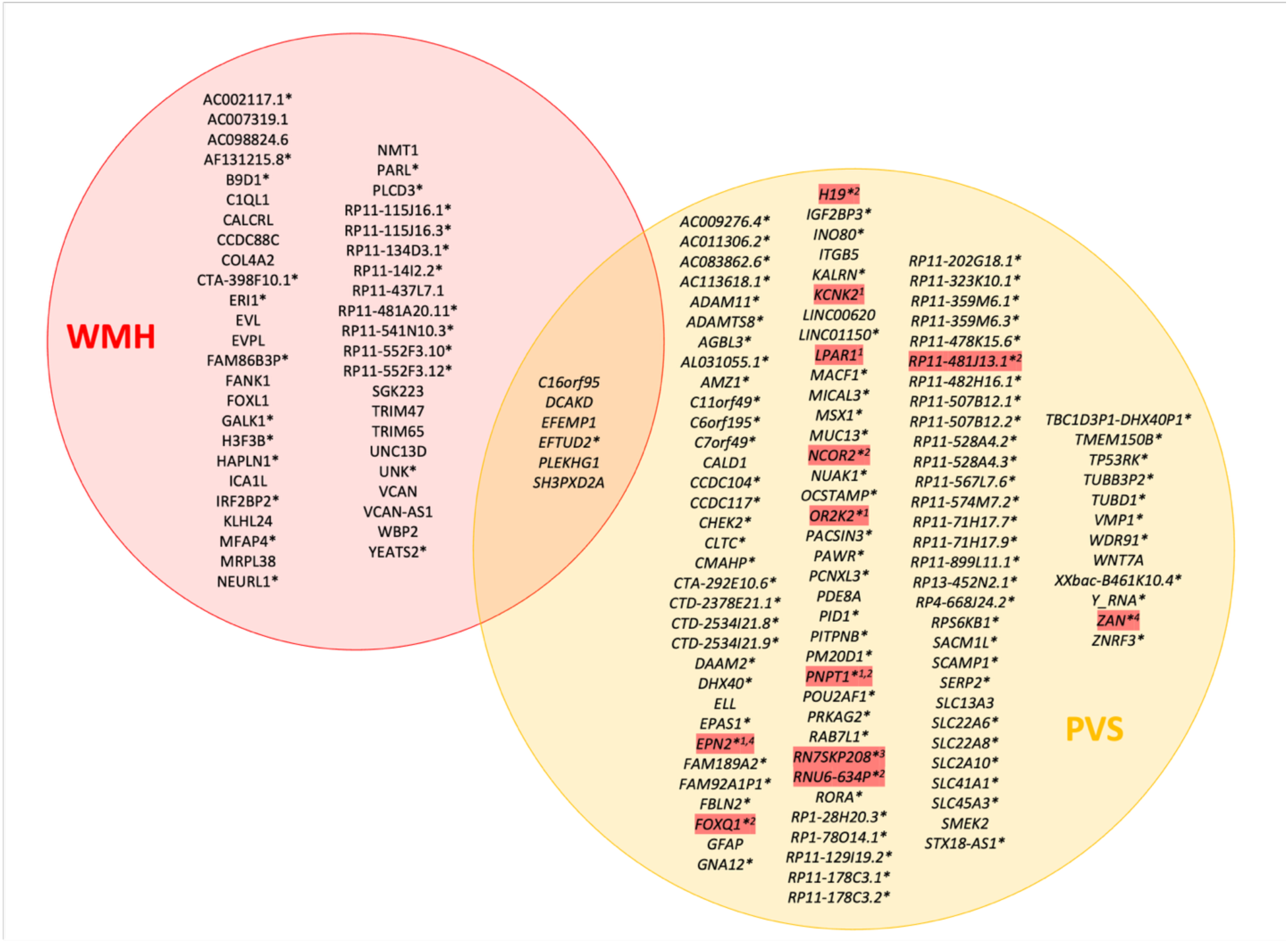
Venn diagram showing genes nearest to independently significant SNPs for PVS volume and WMH volume. *novel genes for respective phenotype; marked in red are genes identified in GWAS on PVS volume, that were previously reported in GWAS on WMH. ^1^ Sargurupremraj M, ^2^ Armstrong NJ,^3^ Rutten-Jacobs LCA, ^4^ Persyn E

In addition to positional mapping, we used eQTL mapping, chromatin interaction mapping, and MAGMA gene-based association analysis to map the identified SNPs to genes. Twenty-three genes were identified by all four methods. The strongest signals were mapped to *ITGB5*, *TUBD1*, *RAB7L1*, *ZNRF3*, *SCAMP1*, *EFEMP1* and *UMPS* (Supplementary Figure 3).

Using FUMA’s GENE2FUNC module, we conducted an enrichment analysis to determine whether the genes tagged by our lead SNPs were significantly enriched in curated gene sets associated with other traits in the GWAS catalogue (Figure 3). This analysis revealed that genes associated with PVS volume in our GWAS were also enriched in gene sets linked to traits, including medial thalamic nuclei volume, WMH volume, PAI-1 levels, Alzheimer’s disease and lacunar stroke. Notably, approximately 55% of the genes identified in our PVS volume GWAS overlapped with genes previously linked to PAI-1.

**Figure 3:**
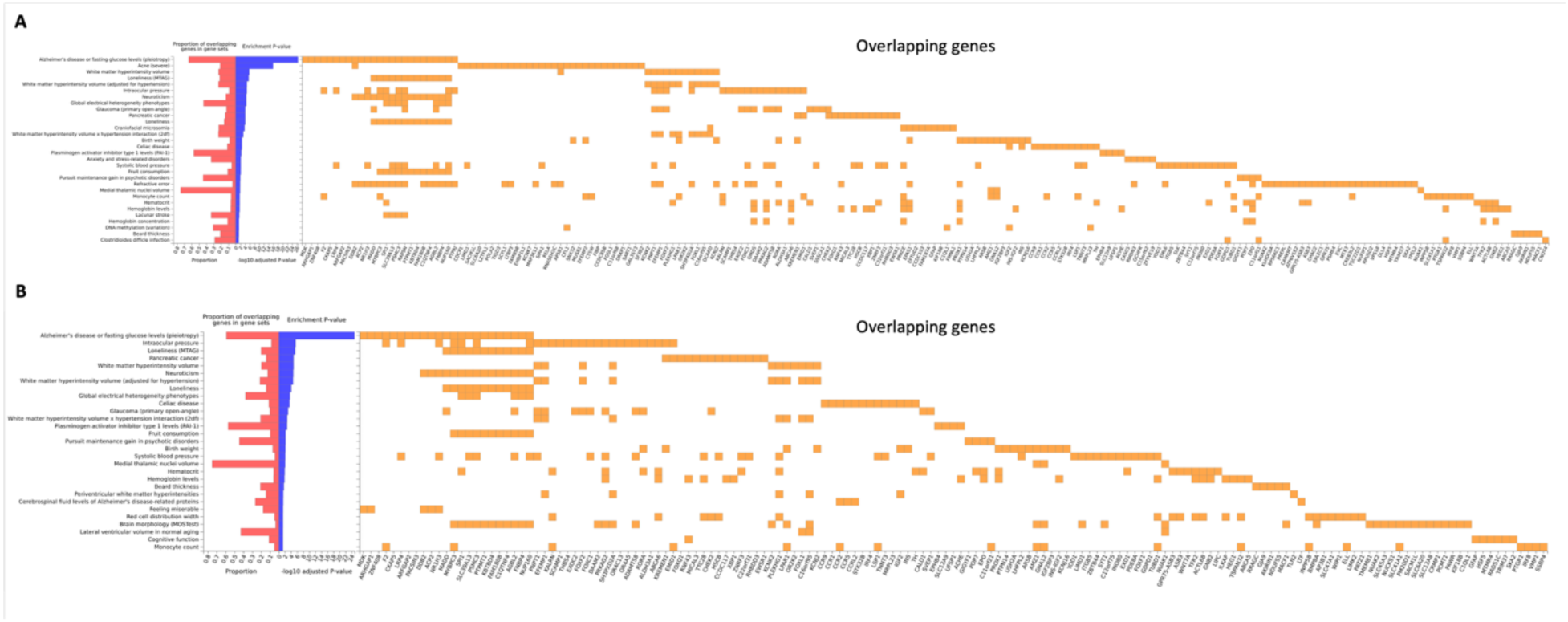
Graphic showing genes identified in GWAS on PVS volume (A: model 1, B: model 2), which are enriched in gene sets derived from prior GWAS studies, curated in the NHGRI-EBI GWAS Catalog.

### Genetic correlation with clinical correlates

We performed genetic correlation analysis between PVS- and WMH volume, within the same sample as well as with regional PVS burden and DTI-ALPS. There was no significant genetic correlation between WMH volume and PVS volume (model 1: r_g_ = 0.07, P_FDR_ = 0.46). Whole brain PVS volume was genetically perfectly correlated with WM PVS burden (model 1: r_g_ = 1.00, P_FDR_ = 3.40×10^−8^^3^). In contrast, whole-brain PVS volumes had moderate correlations with BG PVS burden (model 1: r_g_ = 0.42, P_FDR_ = 1.14×10^−5^) and low correlations with hippocampal PVS burden (model 1: r_g_ = 0.30, P_FDR_ = 0.001). Strikingly, higher PVS volumes were genetically significantly correlated with higher DTI-ALPS indices, indicating a better glymphatic function (model 1: r_g_ = 0.16, P_FDR_ = 0.003) (Figure 4A, Supplementary Table 20).

**Figure 4:**
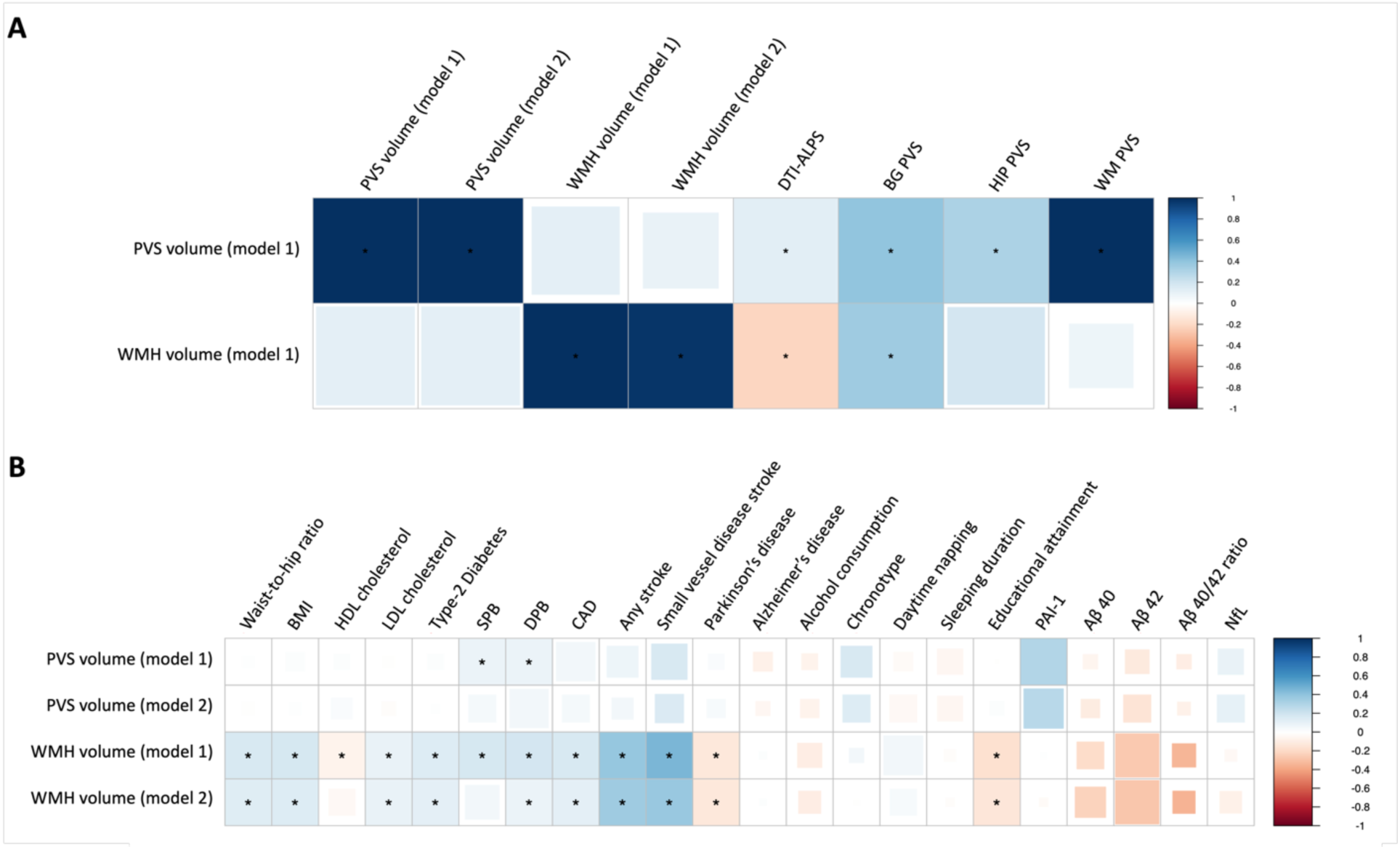
Genetic correlation of PVS volume and regional burden, WMH volume and Diffusion Tensor Imaging - Along the Perivascular Spaces (DTI-ALPS) with each other.

The genetic correlation analysis between PVS and WMH volumes and cSVD risk factors as well as clinical correlates revealed significant genetic correlations of PVS volumes with DBP (model 1: r_g_ = 0.09, P_FDR_ = 0.004) and SBP (model 1: r_g_ = 0.08, P_FDR_ = 0.011). A nominally significant genetic correlation could be observed between PVS volume and PAI-1 (model 1: r_g_ = 0.30, P = 0.02), which did not remain significant after FDR correction (model 1: P_FDR_ = 0.06). Genetic correlations with other vascular risk factors, neurodegenerative diseases and neurological traits, sleep related traits as well as blood-based markers of neurodegeneration were not significant.

In contrast, WMH volume showed significant genetic correlations with vascular risk factors (BMI: r_g_ = 0.17, P_FDR_ = 2.38×10^−10^; WHR: r_g_ = 0.16, P_FDR_ = 6.90×10^−8^, LDL cholesterol: r_g_ = 0.10, P_FDR_ = 0.009; SBP: r_g_ = 0.17, P_FDR_ = 1.32×10^−6^; DBP: r_g_ = 0.19, P_FDR_ = 1.64×10^−8^), clinical outcomes (CAD: r_g_ = 0.17, P_FDR_ = 1.81×10^−5^; any stroke: r_g_ = 0.38, P_FDR_ = 6.55×10^−1^^4^; SVD stroke: r_g_ = 0.45, P_FDR_ = 3.62×10^−5^; Parkinson’s disease: r_g_ = -0.13, P_FDR_ = 0.02) and educational attainment (r_g_ = -0.16, P_FDR_ = 1.30×10^−7^).

### Mendelian randomization with risk factors and clinical outcomes

To explore potential causal relationships, we conducted two-sample MR analyses to investigate associations between key risk factors and PVS volumes, as well as between PVS volumes and clinical outcomes. As candidate risk factors, we focused on SBP and DBP and PAI-1 levels, as these showed the strongest signals in our genetic correlation analyses. Clinical outcomes of interest included stroke, Alzheimer’s disease, and Parkinson’s disease.

To avoid sample overlap, we derived genetic instruments for SBP and DBP from a GWAS conducted in an independent subset of the UKB, not included in the imaging analysis. MR analyses revealed a consistent and statistically significant causal relationship between both SBP and DBP and increased PVS volume across all applied MR methods (Figure 5). These associations remained robust after adjusting for hypertension, reinforcing the link between elevated blood pressure and increased PVS volumes (Figure 5 C, F).

**Figure 5:**
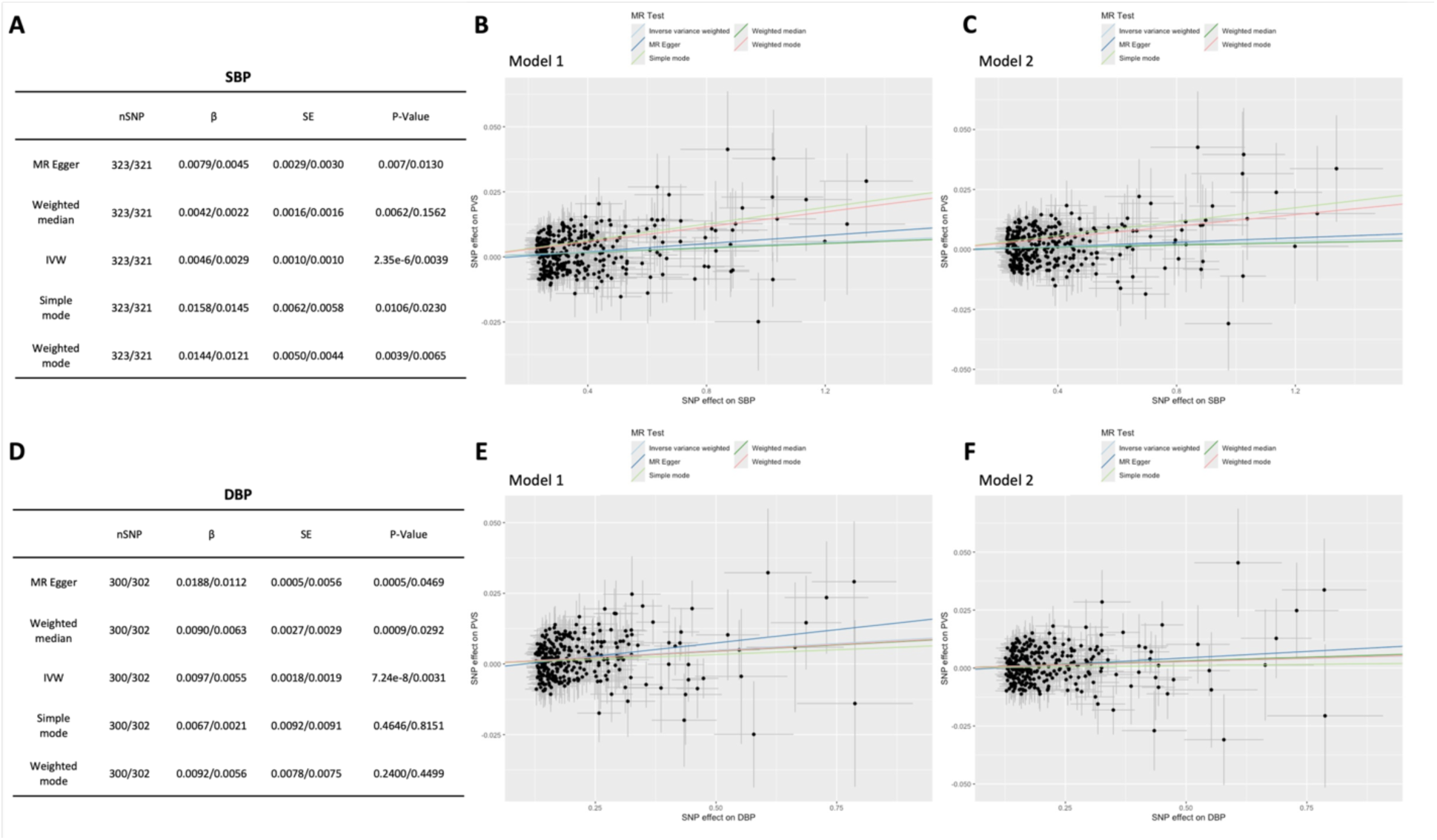
Mendelian randomization (MR) analysis of perivascular space (PVS) volume on systolic (SBP) and diastolic blood pressure (DBP). Panels A and D summarize the causal estimates (β), standard errors (SE), and p-values from five MR methods. Results are shown as Model 1 / Model 2, where Model 1 adjusts for age, sex, estimated total intracranial volume (eTIV), and genetic principal components (PCs), and Model 2 includes additional adjustment for hypertension status. Panels B and C show scatterplots of SNP-exposure vs. SNP-outcome effects for SBP under Models 1 and 2, respectively; Panels E and F show the corresponding plots for DBP. Each point represents a genetic instrument (SNP), and lines indicate fitted effect estimates from inverse-variance weighted (IVW), MR Egger, and other MR methods.

In addition, elevated PAI-1 levels were significantly associated with greater PVS volume in the weighted median MR method (model 2: P_FDR_ = 0.04). Other MR methods, including MR-Egger and mode-based estimators, did not support a significant association. However, this analysis was based on only four SNPs, limiting instrument strength and requiring cautious interpretation. We found no evidence supporting a causal association between PVS volume and Alzheimer’s disease, Parkinson’s disease, or any stroke subtype in the MR framework (Supplementary Table 21).

## Discussion

We identified 54 genome-wide significant risk loci for quantitative measures of PVS volume, 39 of which were not previously reported. Gene-set enrichment pointed to pathways linked to PAI-1, WMH volume, Alzheimer’s disease and lacunar stroke. Genetic correlation analysis confirmed relations to PAI-1 and highlighted shared genetic signatures with systolic and diastolic blood pressure. Interestingly, genetic correlations with WMH indicated largely distinct genetic architectures, challenging assumptions of shared heritability across cSVD markers. Mendelian randomization further demonstrated causal effects of systolic and diastolic blood pressure on PVS burden and supported a role for PAI-1.

Compared to an earlier, and thus far the only reported, GWAS of PVS burden^11^, we find substantially more genetic loci to be associated with PVS. This may have been due to our slightly larger sample size, the state-of-the-art imaging protocols in both the UKB and the Rhineland Study, but above all recent advances in automated PVS segmentation, which allowed us to assess whole-brain PVS quantitively, providing greater power to detect genetic variants linked to PVS enlargement. Even though we used the same automated pipeline to assess PVS in all participants, mean PVS volume and count differed between cohorts, with Rhineland Study participants on average showing larger volumes and higher counts than UKB participants. These differences likely reflect variations in imaging protocols and population demographics, and emphasize the importance of meta-analytic approaches. Despite down-sampling Rhineland Study images, acquisition differences, like specific MRI system, sequence contrast and noise may have influenced PVS quantification. Nevertheless, directions of findings were consistent between cohorts, with 20 loci replicating in the Rhineland Study despite the smaller sample size and stringent correction threshold, underscoring their robustness.

The SNP-based heritability of global PVS volume in our study was 30%, which is considerably higher than previous estimates for regional PVS burden ranging 5-11% for specific regions^11^. This likely reflects advantages of continuous, whole-brain PVS measures, capturing broader biologically relevant variation and improving the power to detect genetic effects. Notably, the 30% heritability is also more in line with reported estimates for WMH, which are 20-30% in SNP-based analyses^5,15^.

In total, we identified 117 different genes associated with PVS volume. Among these genes, *SLC13A3* (tagged by rs847073), emerged as the gene nearest to the most significant locus. *SLC13A3* is part of the solute carrier (SLC) family, which is the largest class of transporters, an important candidate for drug target development^67^ and was already identified as a key PVS risk variant in the previous GWAS on PVS. Here it was proposed that SLCs are involved in interstitial fluid accumulation along PVS, through their role in CSF secretion and substance transport^11^. We identify an additional member of the SLC family, *SLC22A8*, involved in the removal of substrates like estrone sulfate or taurocholate, benzylpenicillin, 2,4- dichlorophenoxyacetic acid and polycyclic aromatic hydrocarbons, from the CSF to the blood^68^, supporting the hypothesis that impaired function of transporters involved in detoxification is also implicated in PVS formation.

Among the novel genetic risk loci, *RORA* (tagged by rs2247306) presented as a highly significant risk variant. The protein encoded by *RORA* is a member of the NR1 subfamily of nuclear hormone receptors. Interestingly, *RORA* has been found to aid in the transcriptional regulation of genes involved in the circadian clock function^69^. The glymphatic system, which clears waste through the perivascular pathways shows increased activity during sleep^10,70,71^. Thus, a disruption of *RORA* might impair sleep patterns, reducing glymphatic efficiency and contributing to PVS enlargement. In addition, *RORA* is a crucial regulator of neurodevelopment, specifically the development of the cerebellum, cellular differentiation and immunity and interacts with genes involved in the regulation of cholesterol and hepatic glucose metabolism^72,73^. This suggests that the relationship between *RORA* and PVS is likely multifactorial. Several of the novel loci identified in this study are functionally linked to vascular development, structural integrity, and remodeling of vessels. For example, *EPAS1* (tagged by rs11125071) upregulates vascular endothelial growth factor, promoting angiogenesis and vascular homeostasis^74^. *ADAMTS8* (tagged by rs7120276) is involved in extracellular matrix degradation and has been implicated in pulmonary arterial hypertension and endothelial dysfunction^75^. Similarly, *MSX1* (tagged by rs75558778) is expressed in vascular smooth muscle cells and pericytes, suggesting a role in vessel wall morphogenesis and stabilization during development and repair^76^. Other novel identified genes are involved in immune regulation and inflammation. *POU2AF1* (tagged by rs11825875) encodes a transcriptional coactivator essential for B-cell function and antibody-mediated immunity^77^, while *H19* (tagged by rs115576433), has been implicated in modulating inflammatory signaling and vascular aging^78^. Collectively, these findings support a multifaceted model in which genetic variation affecting vascular structure, cytoskeletal remodeling, inflammation, and metabolic regulation contributes to PVS enlargement, consistent with prior reviews suggesting a multifactorial etiology for PVS^12,13^.

Of the identified PVS risk loci, 41% emerged as known cSVD marker risk loci, mostly related to WMH and brain atrophy, supporting the hypothesis that PVS are a marker of cSVD. However, the overlap of genes identified in GWAS of PVS volume and GWAS of WMH volume in the same sample was only minimal with 6 out of 163. Additionally, the genetic correlation of PVS and WMH volume in the same sample was not statistically significant. This suggests that, although these brain pathologies might be markers of the same disease, they appear to have unique genetic underpinnings. Additionally, the genetic correlation analysis between PVS- and WMH volume and proposed risk factors as well as clinical outcomes highlighted differences between the two MRI-markers of cSVD. While genetic correlations of WMH volume were significant with anthropometric measures like BMI and waist-to-hip ratio as well as blood pressure measures, genetic correlation coefficients for PVS volumes were highest for DBP even when adjusting for hypertension. These findings align with epidemiological evidence linking higher DBP to greater PVS burden, whereas BMI associates only with BG-PVS^7^.

We performed a supplementary GWAS comparing minimal versus higher PVS burden to explore genetic resilience. Given the near ubiquity of PVS in older adults^8^, individuals with minimal burden may offer valuable insight into protective mechanisms. While this approach identified loci not previously reported in region-specific PVS studies, findings largely overlapped with our main quantitative analysis. Notably, one distinct association near *LAMC1* (1q25.3), previously linked only to hippocampal PVS, emerged in an age-stratified analysis, suggesting potential regional specificity in genetic resilience. This implies that continuous PVS volume measures may be more effective in capturing the underlying genetic architecture than dichotomized phenotypes, while also highlighting the potential of extreme phenotypic contrasts for future resilience-focused research. Recent studies have shown that integrating multiple MRI markers into composite cSVD phenotypes can enhance specificity by capturing a broader spectrum of disease burden^79,80^. Additionally, we examined dominant and recessive models to better capture non-additive effects. The recessive model largely recapitulated findings from the additive GWAS, yet revealed two additional loci in each model variant not identified under the additive assumption. Of particular interest, a signal at 11q13.3, previously associated with white matter PVS burden, was detected in both recessive analyses, suggesting a possible mode-specific contribution to PVS pathogenesis. In line with previous studies, which have reported notable sex differences in PVS prevalence and distribution^9^, sex-interaction analysis revealed *CHL1* effects likely to be specific to women. This gene is known to be involved in nervous system development and synaptic plasticity^81^.

Our two-sample MR analyses revealed causal associations of both SBP and DBP with PVS volume. This association remained significant after adjusting for hypertension, indicating that higher blood pressure increases PVS volume even below clinical thresholds. These findings are in line with previous MR results from large-scale GWAS on WMH, which also reported a causal relationship between elevated blood pressure and WMH beyond clinical thresholds of hypertension^5^. Moreover, our results align with those from the previous GWAS on regional PVS burden, which reported consistent associations of genetically elevated SBP and DBP with PVS in all regions^11^. Notably, here, the association with SBP was only nominally significant for WM-PVS and models did not adjust for hypertension status. By including hypertension in our models, our study extends previous findings and highlights the robust and independent role of blood pressure in PVS enlargement, underscoring vascular dysfunction as a key contributor to cSVD pathology.

The significant enrichment of PVS-associated genes in gene sets linked to PAI-1 measurements, a serpin inhibitor known for its regulation of fibrinolysis, hints at a potential role of vascular inflammation and endothelial dysfunction in PVS formation^50,82^. Complementing this, we observed a nominally significant genetic correlation between PAI-1 and PVS, indicating shared genetic factors may underlie both traits. Strengthening this link, our two-sample MR analysis, indicated a potential causal relationship between elevated PAI-1 levels and increased PVS volume. However, this analysis was based on only four genetic instruments, likely reflecting the limited power of the PAI-1 GWAS due to its relatively small sample size. The reduced number of robust instruments may weaken the reliability of causal inference. Future studies leveraging larger GWAS datasets for PAI-1 could strengthen instrument validity and help clarify its role in PVS enlargement. Despite these limitations, the convergence of findings from gene set enrichment, genetic correlation, and MR analyses supports the biological plausibility of a link between prothrombotic states and early microvascular changes in cSVD.

We would like to point out that our analyses were restricted to participants of European ancestry, reflecting the demographic composition of the included cohorts. As a result, the generalizability of our findings to other ancestral populations is uncertain. Future GWAS of PVS volume in more diverse populations are necessary to assess the transferability of genetic associations across ancestries and to improve the inclusiveness of genetic risk prediction.

In conclusion, we identified 54 genetic risk loci associated with global PVS volume, of which nearly three-quarters were novel, i.e., not previously reported in GWAS of regional or dichotomous PVS measures, underscoring the added power and specificity of quantitative trait analysis. Our results point to a complex, multifactorial genetic architecture underlying PVS enlargement. The consistent evidence for a causal role of both systolic and diastolic blood pressure, along with suggestive associations with PAI-1 levels, further highlights vascular dysfunction as a key biological mechanism. Collectively, these findings advance our understanding of cSVD pathophysiology and offer promising directions for preventive and therapeutic strategies.

## Data Availability

The Rhineland Study dataset is not publicly available because of data protection regulations. Access to data can be provided to scientists in accordance with the Rhineland Study Data Use and Access Policy. Requests for further information or to access the Rhineland Study dataset should be directed to rs-duac{at}dzne.de. All individual level data from the UK Biobank Imaging Study used in the current manuscript are available through the UK Biobank Resource (https://www.ukbiobank.ac.uk). This research has been conducted using the UK Biobank Resource under Application Number 82056.

## Contributors

DF, AMI, MMBB, and NAA conceptualized the project. The first draft of the manuscript was written by DF and AMI. Data were analyzed by DF and AMI. SE, MS, NAA, and MMBB provided technical, statistical and methodological advice. All authors provided critical feedback and contributed to the writing and revision of the final version of the manuscript.

## Acknowledgments

The Rhineland Study is funded by the German Center for Neurodegenerative Diseases (DZNE). This work was further supported in part by the German Federal Ministry of Education and Research (BMBF) through grant [FKZ: 01KX2230] with the title "PreBeDem - Mit Prävention und Behandlung gegen Demenz” and the Helmholtz Association under (ExNet-0008-Phase2-3) and the 2025 Innovation Pool. N. Ahmad Aziz is supported by a European Research Council Starting Grant (Number: 101041677)

